# Estimate of airborne transmission of SARS-CoV-2 using real time tracking of health care workers

**DOI:** 10.1101/2020.07.15.20154567

**Authors:** Bala Hota, Brian Stein, Michael Lin, Alex Tomich, John Segreti, Robert A. Weinstein

**Author notes:** Correspondence to: Bala Hota MD, MPH, VP, Chief Analytics Officer, Associate CMO, Professor, Medicine/Infectious Diseases.

## Abstract

**BACKGROUND:** Whether and to what degree SARS-CoV-2 is spread via the airborne route is unknown. Using data collected from health care worker interactions with hospitalized patients with COVID-19 illness, we calculated the transmissibility of SARS-CoV-2 via the airborne route.

**OBJECTIVES/METHODS:** Healthcare worker interaction with SARS-CoV-2 infected patients were tracked using a real time location system between March 18 and March 31. A value for q, the transmissibility expressed as quanta per hour, was estimated using a well-established model for airborne transmission.

**RESULTS:** SARS-CoV-2 infection prevalence among tracked HCWs was 2.21% (0.07-4.35). Transmissibility was estimated to be 0.225 quanta per hour, well below other well-characterized airborne pathogens. Simulations demonstrated that risk of infection is substantially reduced with increased ventilation of rooms.

**CONCLUSIONS:** Overall, our findings suggest that SARS-CoV-2 is not well transmitted via the airborne route in controlled conditions. We speculate that SARS-CoV-2 may be only opportunistically airborne, with most transmission occurring via droplet methods.

**One Sentence Summary:** We calculated the airborne transmissibility (q) of SARS-CoV-2 and the impact of masks and ventilation in a hospital setting.

## Main Text

Since its emergence in late 2019(*1*), COVID-19 has spread internationally as a major cause of morbidity and mortality, and was declared a pandemic on March 11, 2020(*2*). It has at the time of this writing infected 10.9 million and caused over 520,000 deaths worldwide, and over 2.7 million cases and over 120,000 deaths in the United States. The rapid spread of SARS-CoV-2 in populations is attributable to a published reproductive number (R_o_) that has ranged from 2.0 - 3.58(*3–5*). To slow the spread of SARS-CoV-2 in populations and reduce R_o_, public health authorities have recommended control measures that rely on so-called non-pharmaceutical interventions (NPIs) which include social distancing, home isolation for non-essential workers, hand hygeine, and public use of surgical masks. NPIs have been shown in statistical modeling to reduce the likelihood of overwhelming the healthcare system in the setting of rapid case growth(*6*). In the healthcare setting, engineering and administrative controls are critical factors in reducing risk of transmission along with use of personal protective equipment(PPE)(*7*). In the absence of a vaccine and with areas worldwide with transmission that is not contained, shortages of personal protective equipment (PPE) and ventilators may occur; shortages of appropriate PPE may lead to risks for healthcare workers(*8*).

Nosocomial transmission to healthcare workers of respiratory viruses like SARS-CoV-2 occurs primarily via the droplet and contact routes. Airborne transmission has been describe to occur obligately, preferentially, or opportunistically(*9*). The impact of healthcare worker transmission can be significant: during the SARS CoV-1 epidemic, healthcare workers represented 21% to over 50% of cases based on the country, with associated mortality(*10*), and airborne spread was thought to occur(*11, 12*). Data suggest that influenza, including pandemic flu(*13*), can be transmitted via airborne transmission(*14, 15*), as can common colds(*16*) including rhinovirus(*17*); and measles is a known pathogen with clear airborne transmission risk(*18*). It is likely that some viral respiratory pathogens are opportunistically transmitted via airborne routes. Whether and to what degree SARS-CoV-2 is spread via airborne route, i.e. by droplet nuclei is unknown.

CDC(*19*) and WHO(*20, 21*) guidance report that person-to-person transmission of SARS-CoV-2 is dependent in most cases on close contact with infected individuals and occurs via respiratory droplets and contact with contaminated fomites. Current CDC guidance for healthcare workers (HCWs) in the time of SARS-CoV-2 requires wearing a facemask at all times in healthcare facilities, prioritization of facemasks for HCWs, transmission based precautions when in patient rooms, and use of gowns, eye protection such as masks or goggles, and gloves. CDC guidance also recommends that either N95 respirators or surgical facemasks can be worn in patient rooms based on the availability and supply of N95 masks(*19*). For instances where aerosol producing procedures will be performed, N95 respirators or powered air purifying respirators are recommended. Additionally, patients are preferentially placed in single person rooms with doors closed; use of airborne isolation rooms (ie, rooms that are single patient rooms, with a minimum of 6, but ideally 12, air changes per hour that is either filtered before recirculation or vented to the outside) if aerosol generating procedures will be performed; and consideration of dedication of entire units and HCWs in the facility to COVID-19 care.

An underlying assumption for these recommendations is that SARS-CoV-2 is predominantly transmissible via droplet and contact routes, and that airborne transmission is rare. Factors that influence the likelihood of airborne transmission of a pathogen include viral factors, such as likelihood of formation of droplet nuclei that can suspend in the air, size of pathogen droplets after exhalation from a host, and viability of the pathogen once in the environment; environmental factors that can influence transmissibility include ventilation in the immediate environment, the number of air exchanges in the room, the use of additional factors such as HEPA filtration and UV light use, and the number of susceptible individuals in the environment.

The risk to healthcare workers from exposure to pathogens with airborne modes of transmission has been modeled by the Wells-Riley equation, and was subsequently modified by Gammaitoni and Nucci(*22*). Both models establish the relationship of various factors that impact the likelihood of transmission of an airborne pathogen. Gammaitoni’s modification is of the form:

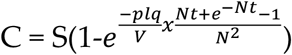

Parameters in the model include the number of new infections (C); the number of susceptible individuals (S); number of infection sources during the exposure event(l); number of doses of pathogen added to the air per unit time (q, in count/hour); pulmonary ventilation per susceptible individual in volume per unit time (p, in *m*^3^/hour), adjusted for the mask scale factor; exposure time (t, in hours); the volume of the environment where infected and susceptible individuals are present (V in *m*^3^); and the total loss/disinfection rate of the environment (N), which is the total aggregate value that represents the net effect of air exchanges, filtration, deposition of particles, inactivation through UV light, and other factors(*23*). The mask scale factor is a measure of mask filtration and is the proportion of airborne nuclei not filtered by the mask and able to be inspired during ventilation. It is calculated as 1 minus the difference of the efficacy of the mask and the product of the face seal leakage of the mask times the mask efficacy. As one example, an N95 mask is 95% effective at filtration of airborne nuclei particles and may have on average 10% leakage. The scaling factor in this case is 1-(0.95 - (0.95*0.10)) = 1-0.855 = 0.145. The scaling factor for perfectly fit N95 masks is 0.05, and for surgical masks is 0.60(*22*).

Assumptions for this model include that exposure risk is homogenous in the environment around the infection source, or patient (i.e. does not vary by distance) and that susceptibility is similar among all contacts between susceptibles and infected individuals.

Of note is the parameter q, which is a measure of the infectiousness via the airborne route of a pathogen. This number represents the number of infectious particles shed by an infectious source per hour into viable droplet nuclei. Values for this number are derived epidemiologically. If the number of infected healthcare workers, the time spent exposed, and room conditions are known, then the value of q can be estimated using the formula:

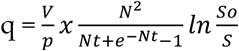

in which S_o_ is the number of infected individuals.

Overall, the Gammaitoni model shows that the net number of infections in susceptible, exposed individuals from an infected source varies directly with amount of virus shed by infected patients and minute volume for breathing of exposed individuals, and inversely with the amount of air cleaning in room via air exchanges, filtration, or pathogen elimination.

Using data collected from healthcare worker interactions with patients with COVID-19 illness, we calculated the transmissibility of SARS-CoV-2 via the airborne route, and evaluated whether SARS-CoV-2 infection rates are consistent with predicted rates of infection due to airborne transmission. Using a retrospective cross-sectional study design, we report overall infection rates among clinical and non-clinical employees. We captured the exact duration of interactions between healthcare workers and patients through the use of real time location services (RTLS) utilized among nursing staff. Using a Monte Carlo simulation applied to these data, we estimated the value of q for SARS-CoV-2, and compared rates of infection through airborne transmission due to SARS-CoV-2 versus other airborne pathogens reported in the literature. Finally, using the estimated value for q, we modeled the probability of infection in individuals exposed to SARS-CoV-2 for multiple scenarios of mask use and room HVAC settings, based on time of exposure.

## Results

### Overall Infection Rates Among Healthcare Workers

Table 1 shows the overall prevalence of infection among healthcare workers at Rush. In the time period, 11,044 employees were observed and followed. Following the algorithm instituted at Rush, employees with symptoms were tested for infection with SARS-CoV-2 following an evaluation by Employee Health, and during this time frame, were self-quarantined following potential exposures. An overall prevalence of 1.62% (95% CI: 1.38-1.86) was noted among all employees; for employees with completely patient-facing roles, the prevalence was 1.85% (1.50-2.20); fully non-clinical roles with no patient-facing contact had a prevalence of 1.11% (0.64-1.58). Rates were not significantly different between the groups.

**Table 1.** Prevalence of infection among various employee groups. Rates reflect testing conducted between March 1 - April 7.

|  | N | Positives | Prevalence | LCI | UCI |
| --- | --- | --- | --- | --- | --- |
| **All Rush Employees** | 11044 | 179 | 1.62% | 1.38% | 1.86% |
| **Non Clinical** | 1888 | 21 | 1.11% | 0.64% | 1.58% |
| **Mixed** | 3589 | 55 | 1.53% | 1.13% | 1.93% |
| **Clinical** | 5567 | 103 | 1.85% | 1.50% | 2.20% |
| **Observed nurse staff** | 181 | 4 | 2.21% | 0.07% | 4.35% |

### Health Care Worker Observations

In the time frame, 2668 healthcare worker-patient interactions were observed among 384 patients. Of these, 152 patients were positive for SARS-CoV-2, with 1198 interactions over 19503 minutes among 181 healthcare workers, for an average of 16.3 minutes per interaction. Of the SARS-CoV-2 positive patients, 63 unique patients were in ICUs on at least one day, and 115 were on general medical floors at least one day. During the period of observation, 4 healthcare workers became infected with SARS-CoV-2, yielding a prevalence of infection of 2.21% (0.07-4.35).

### Observed and Predicted Infection Rates and Estimated Value of q

Parameters used in the Monte Carlo simulation to model the predicted probability of healthcare worker acquisition of SARS-CoV-2 at varying levels of transmission are shown in Table 2. Two scenarios were examined: predominant use of surgical masks by staff; and predominant use of N95s, despite a goal of primarily surgical mask use. In the situation where all healthcare worker interactions occurred with surgical mask use, the value of q is estimated at 0.09 (0.00-0.32). If, on the other hand, healthcare workers were mainly using N95 masks, q is estimated at 0.36 (0.00 - 1.29), or 4 times higher transmissibility via the airborne route, given the better protection afforded by N95 masks for the observed rate of infections.

**Table 2.** Parameters used to calculate q using the Gammaitoni and Nucci model.

| Parameters for Model (per hour) | Value | Units | SD |
| --- | --- | --- | --- |
| p: Pulmonary ventilation per susceptible in volume per unit time. Minimal exertion | 0.48 | m3/hour | 0.12 |
| mask scaling factor (N95) | 0.1 |  |  |
| mask scaling factor (surgical mask) | 0.6 |  |  |
| N: Number of Air Exchanges per Hour | 12 | per hour |  |
| V: Volume of Room | 51 | m3 |  |
| S: Susceptibles | 181 |  |  |
| So: Susceptibles at end of time period | 177 |  |  |
| Infected | 4 |  |  |
| t: exposure time | 325 | hours |  |
| Baseline infection prevalence | 0.011 |  | 0.002 |

Using a mean value for q of 0.225, we calculated estimated prevalence rates of infection for the HCW cohort in various scenarios(Figure 1), including for Rhinovirus (q=5)(*17*), Tuberculosis (q=12.7)(*22*), SARS-CoV-1(q=57)(*24, 25*), and Influenza (q=100)(*17*). Infection rates were significantly higher in the instances of SARS-CoV-1 and Influenza, while TB and Rhinovirus were higher, but not statistically significantly so, compared with SARS-CoV-2 observed and estimated rates.

**Fig 1.**
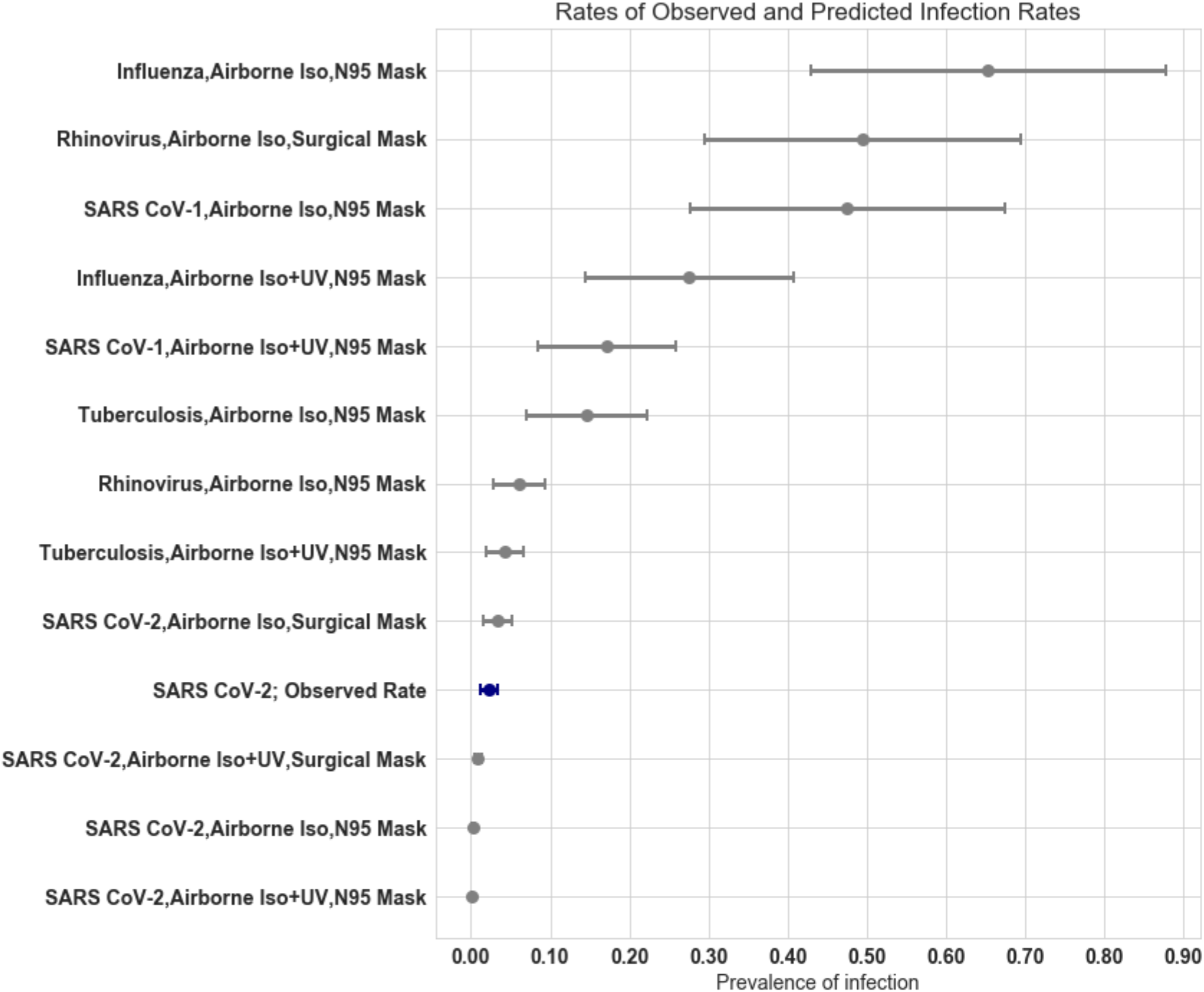
Predicted prevalence based for various pathogens based on observed healthcare worker exposure times. *Values of q for listed pathogens: SARS-CoV-2: 0.225; Tuberculosis: 12.7; Rhinovirus: 5; SARS-CoV-1: 57; Influenza: 100.

### Impact on Expected Infection Rates of Changing Environmental Conditions

The probability of infection of healthcare workers was also modeled with changing duration of exposure, room ventilation, and mask use (Figure 2). Key factors that affect transmission rates are the value for q; the number of effective room air exchanges; and the mask scale factor which is inversely related to mask efficacy. In the models, infection risk increased linearly with longer duration exposures. Risks for up to 48 hours remained low for all room types, while negative pressure rooms with UV light treatment and HEPA filtering had similar rates regardless of mask use, well below 1%.

**Figure 2.**
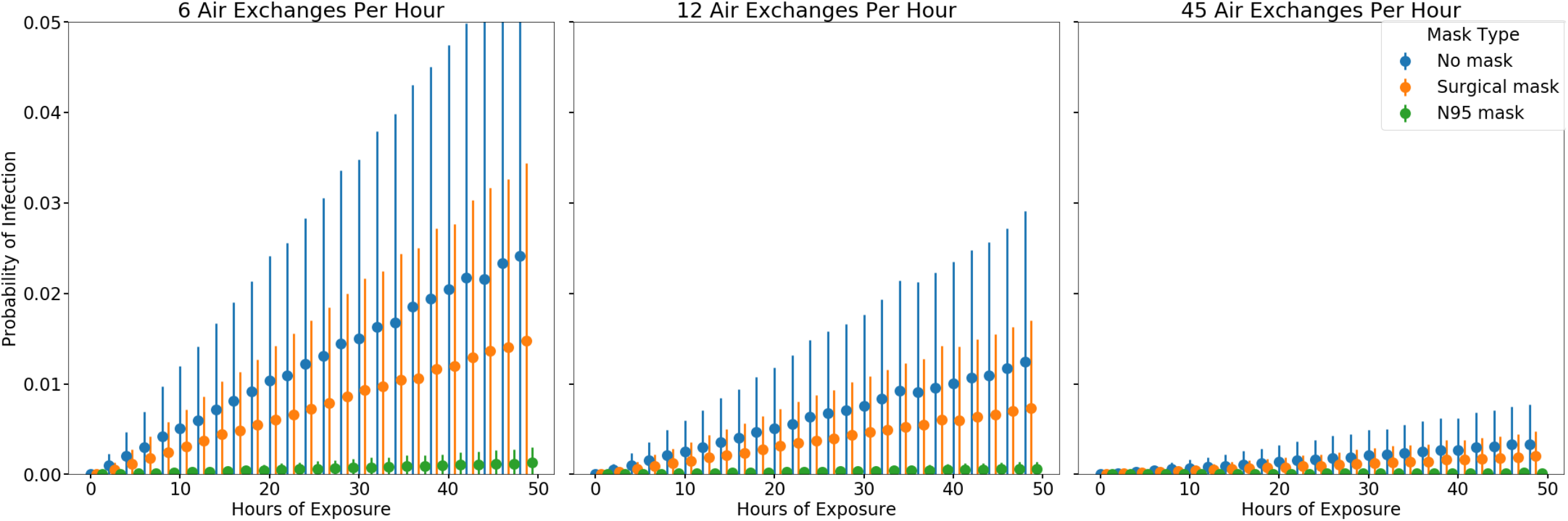
Risk of Infection based on duration of exposure, mask type, and room air exchanges.

## Discussion

Using data obtained from a real time location tracking system and SARS-CoV-2 testing of healthcare workers caring for patients infected with COVID-19, we estimated values for airborne transmissibility for SARS-CoV-2. Our results indicate that the pathogen has lower airborne infection transmission rates than that of Rhinovirus, Tuberculosis, and SARS-CoV-1. We also found that for the estimated value of q for SARS-CoV-2, ventilation controls were most effective in reducing the risk should the pathogen be transmitted by airborne routes. Estimates for q ranged from 0.09-0.37 quanta per hour, with a mean value of 0.225. This value was well below that of other airborne pathogens, including SARS-CoV-1, Tuberculosis, and Influenza. We believe that our study is unique in that it is based on real world data on healthcare worker-patient interactions, and has the benefit of accurate measures of duration of exposure to infected individuals.

A number of aspects of the transmissibility of SARS-CoV-2 are unknown. First, the size of droplets and droplet nuclei from SARS-CoV-2 formed after exhalation have not been established. The formation of droplet nuclei is affected by dehydration of exhaled fluids, which may be affected by humidity and temperature. The viability of the pathogen over time in the environment is also unknown. Data supporting primarily contact and droplet routes for SARS-CoV-2 include low rates of person-to-person spread in initial travel-associated cases of COVID-19(*20, 26*), the absence of virus in airborne samples in an early study from China(*27*) and from Iran(*28*), and a case report of no health care worker conversions after nosocomial exposure to an infected patient(*29*).

In contrast, in a non-peer reviewed report, SARS-CoV-2 was found in air and surface samples in 80% of room surfaces tested, and air samples of hospital rooms and hallways outside the rooms showed 63% and 67% positive rates, respectively(*30*). Additionally, in experimental conditions, SARS-CoV-2 was found to remain viable after aerosolization for 3 hours, similar to SARS-CoV-1. These data may suggest that SARS-CoV-2 can form droplet nuclei and survive for prolonged periods, which are necessary factors to enable airborne transmission. A factor that may affect the duration of aerosolization of droplets in air is the rate at which the droplet will fall from suspension in air, which is directly proportional to droplet size. As droplets are expired, they shrink due to evaporation; larger droplets, which may contain higher quantities of virus, will fall faster than smaller droplets. A recent study found that droplets emitted during spoken phrases can be suspended in air up to 14 minutes, and may be linked with presence of droplet nuclei in some scenarios(*31*).

Additionally, scenarios in which many individuals may be infected from a point source have been described. Examples in which community point source outbreaks have likely occurred due to presumed superspreaders include a choir practice(*32*) and family gatherings(*33*). With the estimate of transmission from the data presented here, superspreader events are likely driven not by the airborne routes, but from closer contact and transmission of virus in respiratory droplets, not droplet nuclei. While the current data could be suggestive of airborne transmission, there continues to be a lack of direct evidence, though the collection of indirect evidence has caused concern(*34, 35*).

The use of modeling is helpful to identify the level of putative transmission via airborne routes, as well as understand the interaction between built environment, the ventilation practices in a room, the use of masks and infectiousness of the source. For SARS-CoV-2, if airborne transmission occurs, it appears that it requires prolonged duration of contact and is effectively reduced in the hospital setting with ventilation controls, consistent with existing strategies for infection control(*7*). Our estimate for the value of q was similar to that of a non-peer reviewed study based on published outbreaks(*36*). We speculate that SARS-CoV-2 may be only opportunistically airborne, with most transmission occurring via droplet methods. For influenza, which also may have mixed features in its transmission patterns, use of surgical and N95 masks have been found to be equally effective in prevention of transmission(*37*). Opportunistic airborne transmission may provide the most reasonable explanation for current real world evidence, and suggest that it is a rare occurrence for SARS-CoV-2(*38*).

Our study is subject to several limitations. First, we do not have detailed data about what types of masks were used by healthcare workers in their interactions with patients. While a policy to use surgical masks was in place, it is possible healthcare workers may have used N95 masks, and we modeled this as well. Second, we could not fully account for acquisition of SARS-CoV-2 infection through community exposures. We did model in our analyses the rates of community onset infection through the use of observed rates among individuals without patient care exposure. We could not account for potential spread among healthcare workers in non-clinical settings such as break rooms in which close contact between individuals may occur(*39*) or superspreader events or aerosol generating procedures. Third, while we measured healthcare worker-patient interactions in this cohort using the RTLS system, exposures among healthcare workers that aren’t tracked using this system (e.g. physicians) could not be assessed. Fourth, these results are applicable to hospitalized patients with COVID-19; risks in the community may be different. Our modeling did not account for the use of masks, especially surgical masks, by infected individuals, though in practice, inpatients did not wear facemasks unless they were outside their private rooms. Fifth, we did not culture asymptomatic people during this time and our estimates of SARS-CoV-2 prevalence may be underestimates among both non-clinical and clinical healthcare workers. Finally, our study does not confirm or refute the presence of airborne transmission by SARS-CoV-2, but rather, provides statistical modeling to guide epidemiological studies and the ability to compare to other airborne pathogens.

Overall, our findings suggest that SARS-CoV-2 is not well transmitted via the airborne route in controlled conditions. Use of the value of transmissibility that we have calculated may be helpful for future epidemiological studies and inform infection control practices.

## Data Availability

Data and code for analyses can be found at: Hota, Bala, Stein, Brian, & Tomich, Alex. (2020). Data for manuscript Estimate of airborne transmission of SARS-CoV-2 using real time tracking of healthcare workers. [Data set]. Zenodo. http://doi.org/10.5281/zenodo.3934582

## Funding

*None*

## Authors contributions

Bala Hota MD MPH

Conceptualization, Methodology, Software, Formal Analysis, Validation, Visualization, Writing-Original Draft, Writing-Review and Editing

Brian Stein MD

Conceptualization, Data Curation, Validation, Writing-Review and Editing

Michael Lin MD

Conceptualization, Validation, Writing-Review and Editing

Alex Tomich DNP

Conceptualization, Data Curation, Validation, Writing-Review and Editing

John Segreti MD

Conceptualization, Validation, Writing-Review and Editing

Robert Weinstein MD

Conceptualization, Validation, Writing-Review and Editing

## Competing interests

Authors declare no competing interests.

## Data and materials availability

Data and code for analyses can be found at: Hota, Bala, Stein, Brian, & Tomich, Alex. (2020). Data for manuscript “Estimate of airborne transmission of SARS-CoV-2 using real time tracking of healthcare workers.” [Data set]. Zenodo. http://doi.org/10.5281/zenodo.3934582

## Supplementary Materials

### Materials and Methods

#### Design

We conducted a retrospective cross-sectional study to evaluate the rates of COVID-19 infection among healthcare workers caring for patients infected with SARS-CoV-2.

#### Setting

This study was conducted at Rush University Medical Center (Rush), a 727 bed acute care hospital in Chicago, IL. The first patient to test positive with SARS-CoV-2 was seen at Rush on March 4, 2020. From March 4 to March 31, 2,463 patients were tested at Rush, 406 tested positive, and 149 were admitted.

### Patients or participants

Nurses caring for patients with COVID-19 infection between March 18 and March 31, 2020.

#### Methods

To detect potential cases of patients infected with SARS-CoV-2, Rush implemented screening criteria based on CDC and local health department guidance. Until March 11, only those patients were screened that had suggestive travel histories to locations with high incidence rates of COVID-19 disease. Testing criteria were subsequently expanded to include patients with influenza like illness. Testing for patients was also permitted based on physician discretion.

In the Emergency department or via direct admissions, persons under investigation (PUIs) for COVID-19 disease were placed in appropriate infection control precautions. From March 4-March 17, PUIs were isolated in airborne precautions and in negative pressure rooms, and healthcare workers used airborne, droplet, and contact precautions for patient care, with an N95 respirator. Consistent with recommendations from the local health department, infection control precautions were changed to droplet and contact precautions with a face mask alone for routine care of patients, and use of N95 respirators only for aerosolizing procedures, on March 18. PPE was made available from a central supply to healthcare workers, and was stored separately from clinical areas. Healthcare workers were required to request PPE with a goal of 100% of staff adoption of the new guidance to use surgical masks except for in the case of aerosolizing procedures. Training teams were deployed throughout the hospital to observe and correct PPE use. Rooms were cleaned daily with a sporicidal agent (hydrogen peroxide/peroxyacetic acid based formulation). When patients were cleared from having COVID-19 environmental services conducted a more enhanced clean. Lastly, a terminal clean was conducted upon patient discharge.

Nurses at Rush University use the Versus/Midmark real time location software (RTLS) badges as a part of routine patient care. This system uses proximity sensing to detect when healthcare workers are in patient rooms, and the data set is annotated with the individual patient and nurse, with the duration of the interaction. This covers patient interactions in the Emergency Department and inpatients. The ED, two general medical floors, and one ICU were dedicated for cohorting COVID-19 infection patients. Using the RTLS data, interactions between healthcare workers and patients were used to estimate the predicted rate of infection and the actual rate of infection. Interactions between nurses and infected patients were collected. Duration of time in the patient room was also used.

After the first case of COVID-19 infection, Rush instituted policies for infection control, monitoring, and furlough for all employees. All healthcare workers with travel to high risk locations outside Chicago were asked to complete a survey, and self-quarantine until contacted by employee health. Consistent with CDC guidance, all workers with symptoms of respiratory illness or fever were required to refrain from working and were tested for SARS-CoV-2 infection within 1 day. All health care worker testing for COVID-19 occurred at Rush.

All data for patient and employee health testing were stored in the electronic record, and a centralized data warehouse. Using these data, a deidentified data set was created for nursing-patient interactions for SARS-CoV-2 infected patients between March 18 and March 31. Duration of patient contact and total interactions with infected patients were collected.

Detection of SARS-CoV-2 RNA for employees and patients was based upon the real-time PCR amplification and detection of genomic RNA obtained from nasopharyngeal swab testing at the Rush microbiology using RealTime SARS-CoV-2 assay (Abbott Laboratories). The limit of detection of the test was 100 virus copies/mL. Performance characteristics for the Abbott Real-Time SARS-CoV-2 assay have been determined by Abbott Laboratories as part of the Emergency Use Authorization(*40*).

Data on testing and rates of positive tests were collected from test results among employees to measure the prevalence of infections. Employees were classified as clinical (roles which were entirely patient facing and actively seeing patients), non clinical (entirely back office functions), and mixed (some direct patient care or work in patient care areas). Non clinical employees and clinical employees without patient care responsibilities began working from home beginning March 6. The prevalence of infection among employees was measured using healthcare worker test positivity rates; binomial confidence intervals were estimated from the prevalence and number of employees.

A value for q, the transmissibility expressed as quanta per hour, was estimated from known values from the data set. Room volume (V) was collected for the standard room size used for COVID-19 patients. The total loss/disinfection rate of the environment (N) was based on the number of room air exchanges per hour for the rooms on the floors in which COVID-19 patients were admitted. A Monte Carlo simulation was run with 10,000 iterations to model transmissibility (q). For each run, values of p and mask efficacy were sampled from a normal distribution, and prevalence of non clinical infection was sampled from a binomial distribution. Susceptible individuals that were infected were estimated as a proportion of individuals infected through healthcare exposure; this was calculated by obtaining the difference between the actual number of infected individuals, and a sampled estimate of non clinical, community acquired infections. Mean and 95% confidence intervals were obtained from the distribution of results from the Monte Carlo simulation.

To compare the predicted rates of infections via the airborne route with benchmarks, predicted rates of nosocomial infection were calculated for other pathogens using baseline parameters from published literature for rhinovirus, influenza, SARS-CoV-1, measles, and tuberculosis. Expected prevalence was calculated by adding the expected infection probability from airborne transmission with the probability of infection from the community, which was obtained from the prevalence of infection from non-clinical staff. Binomial confidence intervals were used for observed prevalences.

A second simulation was conducted to evaluate the impact of room air exchange rate changes, varying levels of PPE protection, and changes in infection transmissibility (q). Based on the first simulation model, an estimate of q was obtained for SARS-CoV-2. The total loss/disinfection rate of the environment (N) was calculated in the simulation as the total number of air exchanges adjusted for other air cleaning methods including UV light and HEPA filtering. Scenarios with air exchange rates of 2, 6 and 12 were used based on CDC recommendations; 45 was used for a room with 12 air exchanges and HEPA filtering plus UV light. 1000 simulations were run for each set of parameters; p, q, and mask efficacy were sampled from a normal distribution. From this simulation, an estimate of the risk of infection based on duration of exposure was calculated.

All analyses were conducted in the anaconda distribution of python 3.7. This study was presented to the IRB and deemed to be exempt from review given that the data were deidentified and did not contain protected health information.

## Notes

### Competing Interest Statement

The authors have declared no competing interest.

### Funding Statement

No external funding was used for the development of this work.

### Author Declarations

Rush University Medical Center Institutional Review Board

